# Anti-integrin αvβ6 Autoantibodies in the Recurrence of Primary Sclerosing Cholangitis Following Liver Transplantation

**DOI:** 10.64898/2026.09.05.26362063

**Authors:** Koki Chikugo, Takeshi Kuwada, Masahiro Shiokawa, Yoshihiro Nishikawa, Hiroyuki Yoshida, Tatsuki Hirai, Nagomi Mankawa, Ryo Ito, Kota Hashimoto, Yuki Mori, Fumioki Toyoda, Ayako Hirata, Kenji Sawada, Takafumi Yanaidani, Masataka Yokode, Yuya Muramoto, Sakiko Ota, Tomonori Hirano, Yuko Sogabe, Nobuyuki Kakiuchi, Tomoaki Matsumori, Yasuhide Takeuchi, Hajime Yamazaki, Takashi Ito, Tsutomu Chiba, Hironori Haga, Etsuro Hatano, Hiroshi Seno

**Affiliations:** Department of Gastroenterology and Hepatology, Kyoto University Graduate School of Medicine, Kyoto, Japan; Department of Gastroenterology, Kobe City Medical Center West Hospital, Kobe, Japan; Department of Diagnostic Pathology, Kyoto University Graduate School of Medicine, Kyoto, Japan; Section of Clinical Epidemiology, Department of Community Medicine, Kyoto University Graduate School of Medicine, Kyoto, Japan; Department of Surgery, Kyoto University Graduate School of Medicine, Kyoto, Japan; Kansai Electric Power Hospital, Osaka, Japan

**Keywords:** primary sclerosing cholangitis, inflammatory bowel disease, autoantibody, liver transplantation, biomarker

## Abstract

**Background:** Prediction and diagnosis of recurrent primary sclerosing cholangitis (rPSC) after liver transplantation remain challenging. We investigated anti-integrin αvβ6 autoantibody titers and their association with rPSC development.

**Methods:** Twenty-three patients with PSC with serial serum samples and liver biopsies after liver transplantation were included. rPSC was diagnosed according to the 2024 Japanese diagnostic criteria. Anti-integrin αvβ6 autoantibody titers were measured by enzyme-linked immunosorbent assays. Fold changes (FCs) in antibody titers were calculated relative to each patient’s pre-transplant baseline, and the maximum FC during post-transplant follow-up was defined as FCmax.

**Results:** Antibody titers significantly decreased at the first postoperative measurement compared with the pre-transplant level. Ten of 23 patients developed rPSC. Three patients whose ulcerative colitis activity changed after transplantation were excluded from further analysis, leaving 20 patients. The maximum absolute antibody titer during post-transplant follow-up was not associated with rPSC development. However, FCmax was significantly higher in patients with rPSC than in those without rPSC (1.381 vs. 0.514, *P* = 0.041), with an AUC of 0.781.

**Conclusions:** Anti-integrin αvβ6 autoantibody titers significantly decreased after liver transplantation for PSC. Post-transplant antibody re-elevation relative to the pre-transplant baseline was significantly associated with the development of rPSC.

## Introduction

Primary sclerosing cholangitis (PSC) is a rare chronic cholestatic liver disease. Although its incidence and prevalence vary geographically, they have been reported to range from 0 to 1.58 cases per 100,000 person-years and from 0 to 31.7 cases per 100,000 population, respectively [1]. In Japan, the prevalence is estimated to be 1.80 per 100,000 population, making PSC considerably less common than in Western countries [2].

Liver transplantation (LT) is currently the only established curative treatment for patients with advanced PSC [3]. However, recurrent PSC (rPSC) develops in approximately 20–30% of transplant recipients and is associated with reduced graft survival and poorer long-term outcomes [3–6]. Despite its clinical importance, diagnosing rPSC after LT remains challenging. Characteristic cholangiographic findings are often absent during the early stages, whereas histological findings overlap with those of rejection and other post-transplant biliary complications. Consequently, diagnosis requires integration of radiological, histopathological, and clinical findings [7–9]. Therefore, disease-specific serum biomarkers for the objective assessment of rPSC are needed.

PSC is characterized by progressive biliary inflammation and fibrosis leading to liver failure. The disease is frequently associated with inflammatory bowel disease (IBD); in Western countries, approximately 70–80% of patients with PSC have concomitant IBD, most commonly ulcerative colitis (UC) [10]. Although biliary epithelial cells are thought to be the primary target of the disease, the underlying mechanisms of PSC remain poorly understood [11].

Anti-integrin αvβ6 autoantibodies have been identified as highly sensitive and specific biomarkers for UC, with antibody titers correlating with disease activity [12, 13]. They also show high diagnostic performance for PSC regardless of concomitant IBD, as demonstrated in both an initial study and an independent nationwide Japanese cohort [14, 15].

Integrin αvβ6 is an epithelial-specific integrin expressed on biliary and intestinal epithelial cells [14]. It binds fibronectin and the latency-associated peptide (LAP) of latent transforming growth factor-β (TGF-β) and activates latent TGF-β, thereby regulating epithelial homeostasis, barrier function, and immune responses [16, 17]. A patient with a homozygous germline *ITGB6* mutation developed lethal cholestatic liver disease and bloody diarrhea resembling PSC and UC, respectively [18]. Moreover, IgG isolated from patients with UC or PSC inhibits the binding of integrin αvβ6 to fibronectin in vitro [12, 14]. Together, these findings provide biological support for investigating anti-integrin αvβ6 autoantibodies as disease-associated biomarkers.

LT provides a unique opportunity to investigate longitudinal biomarker dynamics because the diseased liver is removed and PSC may subsequently recur in the graft. A previous pediatric study suggested that anti-integrin αvβ6 autoantibody levels decrease after successful LT and may increase in association with the development of rPSC [19]. However, the dynamics of these autoantibodies have not been systematically investigated in adult liver transplant recipients, and their associations with rPSC and histopathological findings remain unclear.

Therefore, we investigated changes in anti-integrin αvβ6 autoantibody titers following LT in adult patients with PSC and evaluated whether these changes were associated with rPSC and histopathological findings, thereby assessing their potential utility as serum biomarkers for monitoring the post-transplant disease course of PSC.

## Methods

### Study Population

Adult patients (≥18 years) who underwent LT for PSC at Kyoto University Hospital between August 2017 and January 2026 were retrospectively screened. Patients with preserved pre- and post-transplant serum samples, who underwent at least one liver biopsy during follow-up, and who had a minimum postoperative follow-up period of 90 days were considered eligible. Of the 25 patients initially identified, two were excluded because their postoperative follow-up periods were shorter than 90 days, leaving 23 patients in the study cohort (Fig. 1). rPSC was assessed through June 8, 2026. Post-transplant changes in UC activity were assessed retrospectively based on clinical records, including gastrointestinal symptoms, changes in treatment, and endoscopic findings when available. To minimize biological confounding, patients with worsening UC after LT or newly diagnosed UC during follow-up were not included in the analyses of associations with the development of rPSC and with histopathology. Consequently, these analyses were performed in the remaining 20 patients (Fig. 1).

**Fig. 1.**
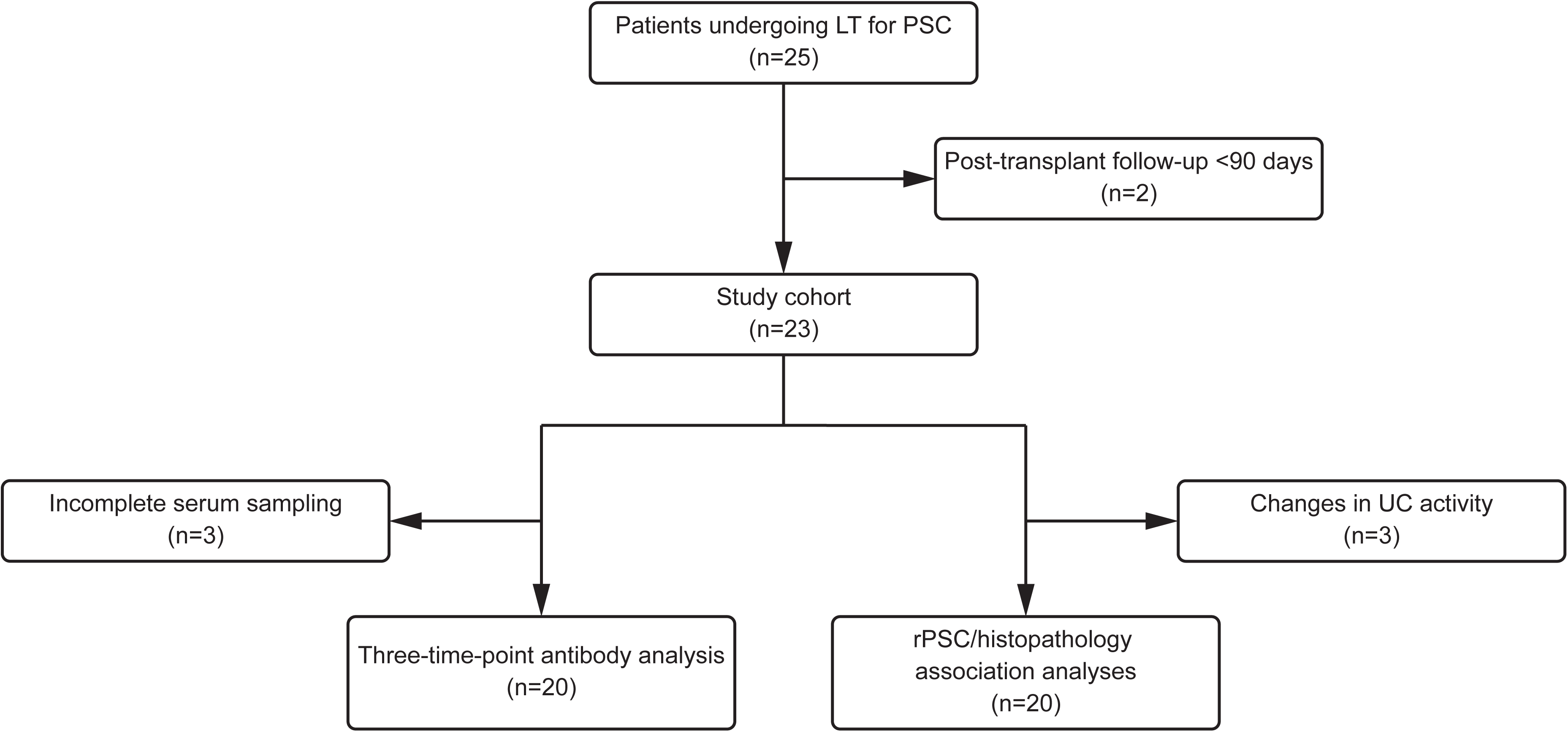
Flow diagram of patient selection and analysis cohorts. Twenty-five patients who underwent liver transplantation (LT) for primary sclerosing cholangitis (PSC) were screened. After excluding two patients with post-transplant follow-up <90 days, 23 patients comprised the study cohort. The three-time-point antibody analysis included 20 patients with serum samples available at the pre-transplant, early postoperative, and postoperative day 90 time points. The rPSC/histopathology association analyses included 20 patients after excluding three patients with post-transplant changes in ulcerative colitis activity. LT, liver transplantation; PSC, primary sclerosing cholangitis; UC, ulcerative colitis

This study was conducted in accordance with the Declaration of Helsinki and was approved by the Ethics Committee of the Graduate School of Medicine and Faculty of Medicine, Kyoto University (protocol number: R1004). Written informed consent was obtained from all participants before study enrollment. Information regarding the study was disclosed on the hospital website, and patients were informed that they could withdraw from participation at any time by contacting the study investigators. This information was also provided directly to the patients.

## Definition of rPSC

rPSC was diagnosed according to the 2024 diagnostic criteria for rPSC after LT [7]. Pre-transplant PSC was histopathologically confirmed in all explanted livers. rPSC was defined by characteristic cholangiographic and/or histopathological findings identified at least 90 days after transplantation after exclusion of hepatic artery thrombosis, rejection, biliary anastomotic complications, and antibody-mediated rejection. Patients classified as Definitive or Probable were considered to have rPSC, whereas those classified as Possible were included in the non-rPSC group.

## Serum Samples

Serum samples were collected between January 2017 and May 2026 during routine clinical care from the pre-transplant period through postoperative follow-up and stored at −80°C until analysis. Samples were generally obtained every few weeks during hospitalization and every 1–6 months at routine outpatient visits after discharge, with additional samples collected as clinically indicated; all available serial samples were included in longitudinal analyses. Because pre-transplant sample availability varied among patients, the sample collected closest to LT was defined as the pre-transplant baseline for all fold-change analyses. The first postoperative serum sample obtained after LT was defined as the early post-LT time point. For analyses at postoperative days 90 and 180, the serum sample collected closest to each target day within a ±30-day window was used.

## Liver Biopsy Specimens and Serum Matching

We retrospectively analyzed liver biopsy specimens obtained during routine clinical practice after transplantation. A total of 84 liver biopsies were performed in the 23-patient study cohort. For each biopsy, the stored serum sample collected near the biopsy date was paired with the corresponding biopsy specimen, yielding 82 matched biopsy–serum pairs for histopathological and antibody analyses.

## Measurement of Serum Anti-Integrin αvβ6 Autoantibodies

Serum anti-integrin αvβ6 autoantibody titers were measured using a commercially available anti-integrin αvβ6 ELISA kit (Catalog No. 5288; Medical and Biological Laboratories, Nagoya, Japan) [15] according to the manufacturer’s instructions. Antibody concentrations were determined using a four-parameter logistic calibration curve generated from serial dilutions of a recombinant human anti-integrin αvβ6 monoclonal antibody standard. Samples with antibody concentrations below the lower limit of quantification were assigned a value of 0.01 U/mL.

## Histological Assessment

Two expert hepatobiliary pathologists independently reviewed all available liver biopsy specimens retrospectively. Whole-slide images of hematoxylin and eosin (H&E)-stained, Elastica-Masson (EM)-stained, and cytokeratin 7 (CK7) immunohistochemically stained sections were generated using a NanoZoomer scanner and assessed for histopathological findings. CK7 immunostaining was not evaluated in two biopsy specimens from one patient because the referring hospital submitted only H&E- and EM-stained glass slides for review. CK7 immunostaining was performed using a ready-to-use mouse monoclonal antibody (Agilent; GA61961-2) on the Dako Omnis platform. Only clinical information necessary for pathological interpretation (e.g., time since transplantation, biliary complications, and vascular complications) was provided to the reviewers. The pathologists remained blinded to antibody titers, rPSC diagnosis, and all study outcomes.

Based on previously reported histological features of biliary injury in PSC and post-transplant cholestatic liver disease [20, 21], three features were evaluated for each biopsy specimen: periductal fibrosis, neutrophilic and/or lymphocytic cholangitis, and periportal CK7-positive metaplastic cells. These features were independently scored using the following semiquantitative scale: 0 (absent), 1 (present in one portal tract), and 2 (present in two or more portal tracts). When both pathologists assigned the same score, that value was used as the final histological score; when scores differed, the mean of the two scores was used. Interobserver agreement for each histological feature was assessed using the weighted kappa coefficient. These histological scores were used to evaluate associations with antibody titers. For the evaluation of T-cell-mediated rejection, the Rejection Activity Index (RAI), as defined by the Banff criteria [22], was obtained from the original pathology report at the time of biopsy without re-evaluation.

**Statistical Analysis**

All statistical analyses were performed using R version 4.6.0 (R Foundation for Statistical Computing, Vienna, Austria). A two-sided *P* < 0.05 was considered statistically significant. Absolute antibody titers were analyzed after log10 transformation when used as continuous outcome variables. Fold changes (FCs) in antibody titers were calculated relative to each patient’s pre-transplant baseline. FCearly was defined as the FC at the first postoperative measurement. FC90 and FC180 were defined as the FCs at postoperative days 90 and 180, respectively. The maximum log10-transformed antibody titer was defined as the highest value observed during follow-up (up to rPSC diagnosis in the rPSC group or the last follow-up in the non-rPSC group). FCmax was defined as the maximum FC observed during follow-up (up to rPSC diagnosis in the rPSC group or the last follow-up in the non-rPSC group), whereas FCbiopsy was defined as the FC at the time of liver biopsy. In an exploratory longitudinal analysis, the first antibody re-elevation above the pre-transplant level was defined as the first postoperative serum sample at which the antibody titer exceeded the individual pre-transplant baseline (fold change >1.0) during follow-up (up to rPSC diagnosis in the rPSC group or the last follow-up in the non-rPSC group). The interval between the first antibody re-elevation above the pre-transplant level and rPSC diagnosis was calculated as the lead time.

Continuous variables are presented as the median (range) or median (interquartile range), and categorical variables as number (%). Comparisons between the rPSC and non-rPSC groups were performed using the Mann–Whitney *U* test and Fisher’s exact test for continuous and categorical variables, respectively. Pairwise comparisons of antibody titers between time points were performed using the Wilcoxon signed-rank test with Holm correction for multiple testing. Receiver operating characteristic (ROC) analyses were performed to evaluate the discriminative performance of antibody-derived variables for rPSC, and the optimal cutoff value was determined using the Youden index.

Interobserver agreement between the two pathologists was assessed using quadratic weighted Cohen’s kappa coefficients, as all three histopathological features independently scored by the two pathologists were evaluated on an ordinal three-point scale (0–2). Associations between histopathological findings and antibody levels were evaluated using linear mixed-effects models with histopathological scores treated as categorical explanatory variables and patient ID included as a random intercept. Two separate linear mixed-effects models were fitted, with the log10-transformed antibody titer at the time of liver biopsy and the fold-change in antibody titer relative to the pre-transplant level (FCbiopsy) as the outcomes in the first and second models, respectively. Regression coefficients (β), 95% confidence intervals (CIs), and two-sided *P* values were estimated for each histopathological category relative to the reference category. Given the exploratory nature of the histopathological analyses, *P* values were not adjusted for multiple comparisons.

## Results

### Patient Characteristics

This study included 23 patients (Fig. 1), whose demographic and clinical characteristics are summarized in Table 1. According to the 2024 diagnostic criteria for rPSC, 10 patients were diagnosed with rPSC. Of these, 2 were classified as Definitive, fulfilling both characteristic histopathological and cholangiographic criteria, whereas 8 were classified as Probable, exhibiting characteristic histopathological findings in the absence of characteristic cholangiographic findings.

**Table 1.** Patient and transplant characteristics according to rPSC status. Continuous variables are presented as median (range), and categorical variables as number (%). LT, liver transplantation; IBD, inflammatory bowel disease; UC, ulcerative colitis; Tac, tacrolimus; MMF, mycophenolate mofetil; EVR, everolimus; PSL, prednisolone; LDLT, living donor liver transplantation; DDLT, deceased donor liver transplantation.

| Variable | Total | rPSC | non-rPSC | *P* value |
| --- | --- | --- | --- | --- |
| Patients, n | 23 | 10 | 13 |  |
| Age at PSC diagnosis, years, median (range) | 28.6 (18.3–64.8) | 31.8 (18.8–45.2) | 28.2 (18.3–64.8) | 0.832 |
| Age at LT, years, median (range) | 41.0 (20.0–69.0) | 44.5 (23.0–55.0) | 41.0 (20.0–69.0) | 0.680 |
| Follow-up duration after LT, years, median<br>(range) | 4.5 (0.6–8.8) | 4.8 (2.0–8.4) | 4.2 (0.6–8.8) | 0.232 |
| Time from LT to rPSC, years, median<br>(range) | 2.3 (0.8–5.7) | 2.3 (0.8–5.7) | - | - |
| Sex, n (%) |  |  |  | 0.685 |
| Male | 15 (65.2) | 6 (60.0) | 9 (69.2) |  |
| Female | 8 (34.8) | 4 (40.0) | 4 (30.8) |  |
| Concomitant IBD, n (%) |  |  |  | 0.379 |
| UC | 15 (65.2) | 8 (80.0) | 7 (53.8) |  |
| No IBD | 8 (34.8) | 2 (20.0) | 6 (46.2) |  |
| New-onset or exacerbation of UC after LT, n (%) |  |  |  | 0.560 |
| Yes | 3 (13.0) | 2 (20.0) | 1 (7.7) |  |
| Cholangiocarcinoma at the time of LT, n (%) |  |  |  | 1.000 |
| Yes | 3 (13.0) | 1 (10.0) | 2 (15.4) |  |
| Immunosuppressive regimen at rPSC diagnosis or last follow-up, n (%) |  |  |  | 0.288 |
| Tac | 1 (4.3) | 0 (0.0) | 1 (7.7) |  |
| Tac + MMF | 5 (21.7) | 4 (40.0) | 1 (7.7) |  |
| Tac + EVR | 1 (4.3) | 1 (10.0) | 0 (0.0) |  |
| Tac + MMF + PSL | 11 (47.8) | 4 (40.0) | 7 (53.8) |  |
| Tac + EVR + PSL | 4 (17.4) | 1 (10.0) | 3 (23.1) |  |
| Tac + MMF + EVR + PSL | 1 (4.3) | 0 (0.0) | 1 (7.7) |  |
| Type of LT, n (%) |  |  |  | 1.000 |
| LDLT | 19 (82.6) | 8 (80.0) | 11 (84.6) |  |
| DDLT | 4 (17.4) | 2 (20.0) | 2 (15.4) |  |
| Intraoperative blood loss, g, median (range) | 2700.0 (417.0–14572.0) | 4533.5 (417.0–14572.0) | 2310.0 (423.0–8640.0) | 0.352 |
| ABO-incompatible LT, n (%) |  |  |  | 1.000 |
| Incompatible | 2 (8.7) | 1 (10.0) | 1 (7.7) |  |

In the study cohort, the median age at PSC diagnosis, age at liver transplantation (LT), and post-transplant follow-up period were 28.6 years, 41.0 years, and 4.5 years, respectively. None of these variables differed significantly between the rPSC and non-rPSC groups. The median time to recurrence in the rPSC group was 2.3 years.

Likewise, no significant differences were observed between the rPSC and non-rPSC groups in sex, donor type, ABO incompatibility, the prevalence of inflammatory bowel disease (IBD), or immunosuppressive regimens.

Fifteen of the 23 patients (65.2%) had concomitant IBD, all of whom had ulcerative colitis (UC); no patients had concomitant Crohn’s disease. During post-transplant follow-up, changes in IBD disease activity were observed in three patients (PSC03, PSC04, and PSC14; 13% of the total cohort and 20% of those with concomitant IBD). Two patients experienced a relapse of UC, while one was newly diagnosed with UC during follow-up. The frequency of post-transplant changes in UC activity did not differ significantly between the rPSC and non-rPSC groups.

### Changes in Anti-Integrin αvβ6 Autoantibody Titers After Liver Transplantation

The pre-transplant and first post-transplant serum samples were collected a median of 4 days (IQR, 2–6 days) before and 7 days (IQR, 5–12 days) after transplantation, respectively. Detailed sampling intervals are summarized in Supplementary Table S1. In 20 of the 23 patients with serum samples available at all three time points (pre-transplant, the first postoperative, and postoperative day 90) (Fig. 1 and Supplementary Table S1), anti-integrin αvβ6 autoantibody titers significantly decreased at the first postoperative measurement compared with the pre-transplant baseline (Wilcoxon signed-rank test with Holm correction, *P* < 0.001). Antibody titers remained significantly lower than pre-transplant levels at postoperative day 90 (Holm-adjusted *P* < 0.001) (Fig. 2).

**Fig. 2.**
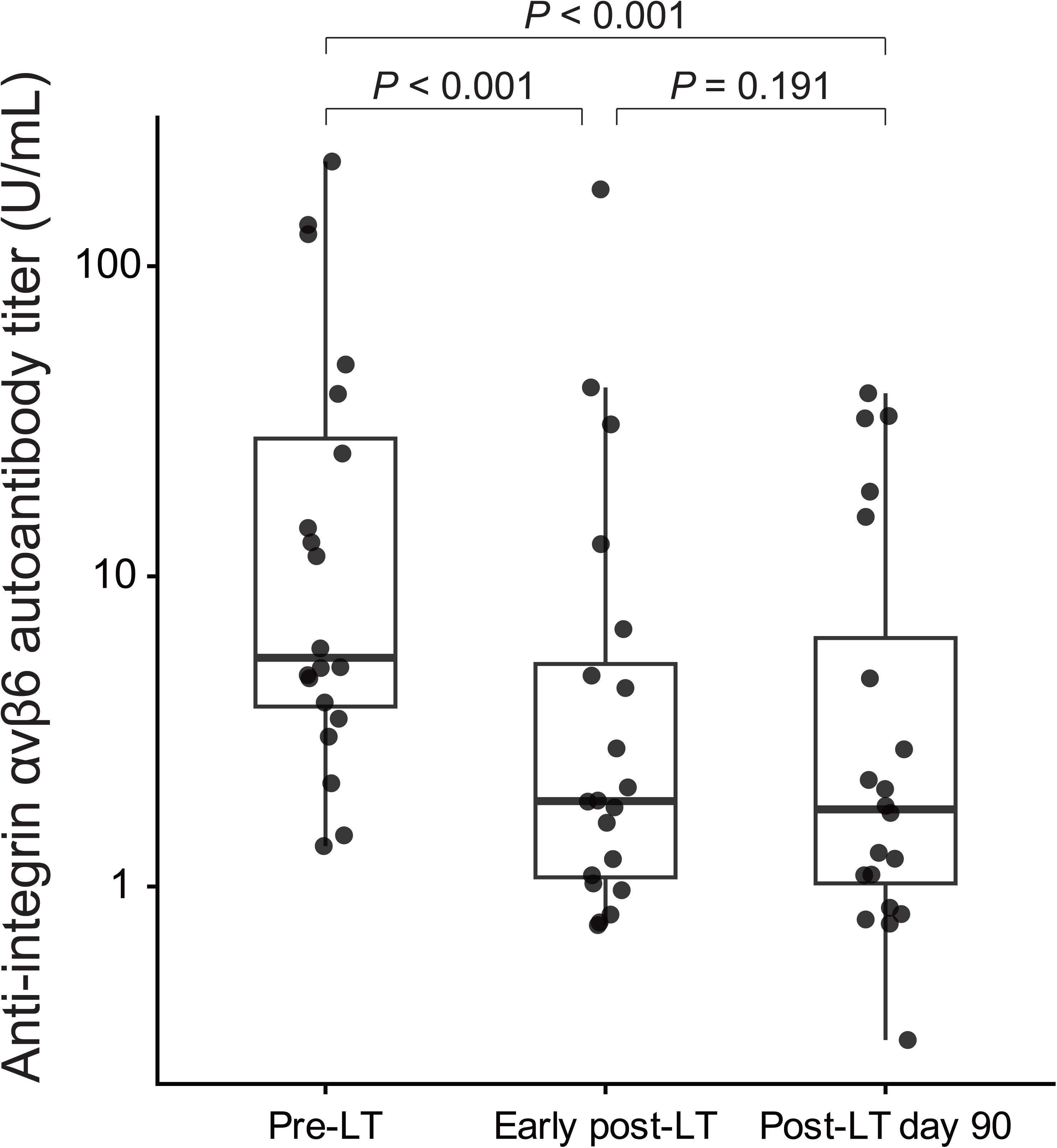
Early postoperative decrease in anti-integrin αvβ6 autoantibody titers after liver transplantation. Anti-integrin αvβ6 autoantibody titers at the pre-transplant (Pre-LT), first postoperative (Early post-LT), and postoperative day 90 (Post-LT day 90) time points in the 20 patients with serum samples available at all three predefined time points. Boxes represent the median and interquartile range, and dots represent individual patients. Pairwise comparisons were performed using the Wilcoxon signed-rank test with Holm correction. Holm-adjusted *P* values are shown

### Re-elevation of Anti-Integrin αvβ6 Autoantibodies During Follow-up After Transplantation

Longitudinal antibody kinetics varied substantially among patients. Fig. 3a and b show representative antibody trajectories for the rPSC and non-rPSC groups, respectively; Supplementary Fig. S1 shows longitudinal antibody profiles for all patients. Pre-transplant anti-integrin αvβ6 autoantibody titers varied considerably among patients, ranging from a few units to over 100 U/mL.

**Fig. 3.**
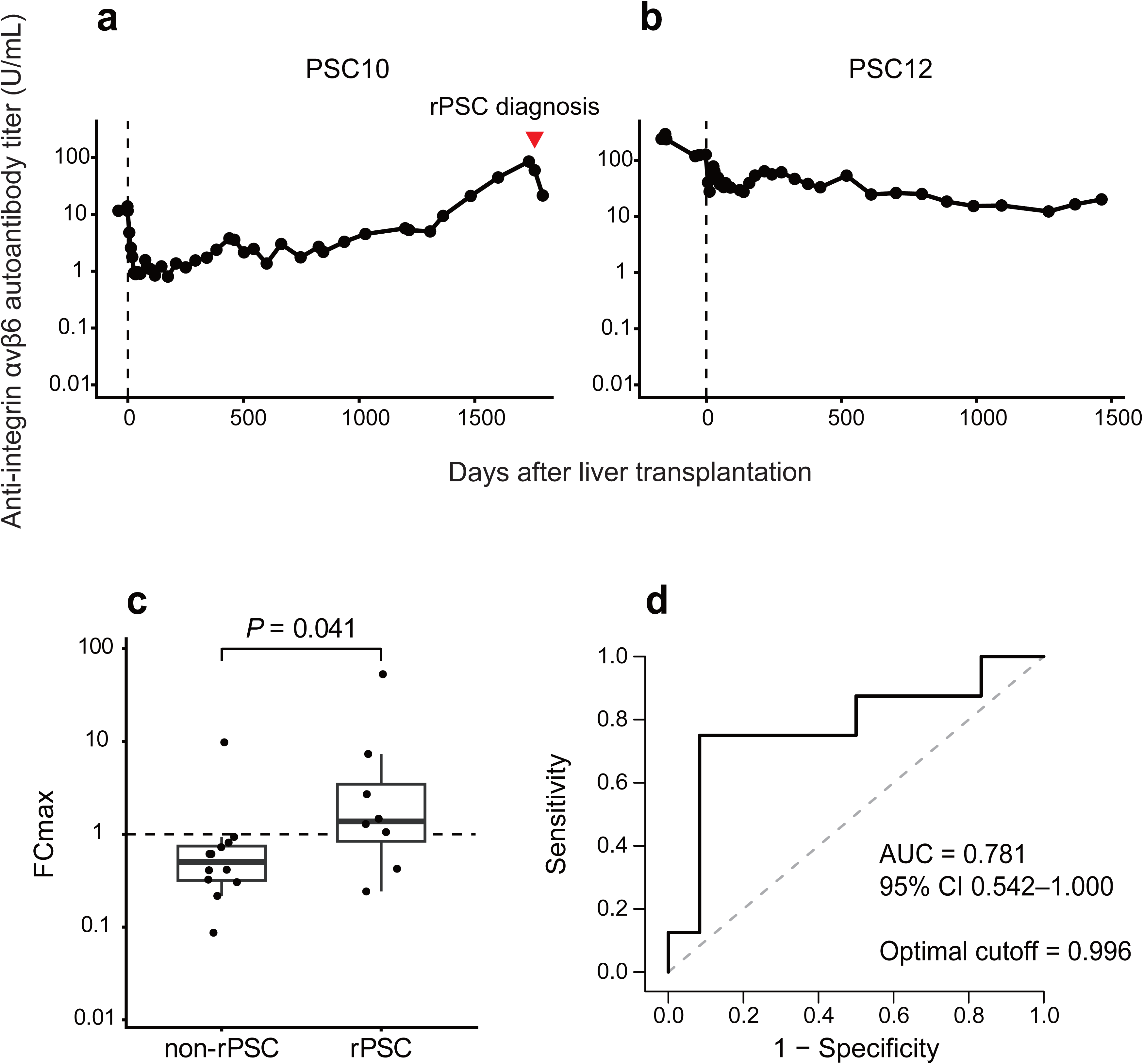
Longitudinal antibody re-elevation and its association with rPSC. a Representative patient with rPSC. b Representative patient without rPSC. Red inverted triangles indicate rPSC diagnosis. The dashed vertical line indicates the time of liver transplantation (day 0). c Comparison of the maximum fold-change in anti-integrin αvβ6 autoantibody titer relative to the pre-transplant baseline (FCmax) between patients with and without rPSC. The dashed horizontal line in panel c indicates an FC of 1.0, corresponding to the pre-transplant baseline. Boxes represent the median and interquartile range, and dots represent individual patients. *P* values were calculated using the Mann–Whitney *U* test. d ROC curve of FCmax for discriminating rPSC from non-rPSC

Table 2 summarizes the longitudinal changes in anti-integrin αvβ6 autoantibody titers, the occurrence of rPSC, and changes in UC activity for each patient. The anti-integrin αvβ6 autoantibody titer in the serum sample collected closest to LT was used as the pre-transplant baseline. During follow-up after transplantation, anti-integrin αvβ6 autoantibody titers increased in several patients. As an initial descriptive assessment, postoperative antibody titers exceeded the pre-transplant baseline at least once in 10 of the 23 patients (43.5%). Three of these 10 patients had post-transplant changes in UC activity, with antibody titers increasing in parallel with UC activity (Supplementary Fig. S1).

**Table 2.** Individual patient characteristics and rPSC diagnostic classification. "Antibody exceeded pre-transplant level" indicates antibody titers exceeding the pre-transplant baseline during follow-up. IBD, inflammatory bowel disease; UC, ulcerative colitis; PSC, primary sclerosing cholangitis; rPSC, recurrent primary sclerosing cholangitis.

| ID | Concomitant IBD | rPSC diagnostic category | Histology compatible with PSC | Imaging findings compatible with PSC | Antibody exceeded pre-transplant level | UC activity change |
| --- | --- | --- | --- | --- | --- | --- |
| PSC01 | No IBD | No recurrence | No | No | No | No |
| PSC02 | UC | No recurrence | No | No | No | No |
| PSC03 | UC | Probable | Yes | No | Yes | Yes |
| PSC04 | UC | Probable | Yes | No | Yes | Yes |
| PSC05 | No IBD | No recurrence | No | No | No | No |

| ID | Concomitant IBD | rPSC diagnostic category | Histology compatible with<br>PSC | Imaging findings compatible with<br>PSC | Antibody exceeded pre-transplant level | UC activity change |
| --- | --- | --- | --- | --- | --- | --- |
| PSC06 | UC | Probable | Yes | No | No | No |
| PSC07 | UC | No recurrence | No | No | No | No |
| PSC08 | UC | Probable | Yes | No | Yes | No |
| PSC09 | No IBD | No recurrence | No | No | Yes | No |
| PSC10 | No IBD | Probable | Yes | No | Yes | No |
| PSC11 | UC | Definitive | Yes | Yes | Yes | No |
| PSC12 | UC | No recurrence | No | No | No | No |
| PSC13 | UC | Definitive | Yes | Yes | Yes | No |
| PSC14 | UC | No recurrence | No | No | Yes | Yes |
| PSC15 | UC | Probable | Yes | No | Yes | No |
| PSC16 | UC | Probable | Yes | No | No | No |
| PSC17 | UC | No recurrence | No | No | No | No |
| PSC18 | No IBD | Probable | Yes | No | Yes | No |
| PSC19 | No IBD | No recurrence | No | No | No | No |
| PSC20 | No IBD | No recurrence | No | No | No | No |
| PSC21 | UC | No recurrence | No | No | No | No |
| PSC22 | UC | No recurrence | No | No | No | No |
| PSC23 | No IBD | No recurrence | No | No | No | No |

### Association Between Antibody Titers and rPSC

Next, we examined the association between anti-integrin αvβ6 autoantibody titers and the development of rPSC. To minimize the influence of UC disease activity on antibody titers, patients with post-transplant changes in UC activity, including UC exacerbation or new-onset UC, were not included in the analyses of associations with rPSC.

Accordingly, the following analyses were performed in 20 patients, comprising those without IBD and those with concomitant IBD without documented post-transplant changes in disease activity (Fig. 1 and Supplementary Table S1).

The median (IQR) of the maximum log10-transformed antibody titer during the post-transplant follow-up period was 0.827 (0.272–1.828) in the rPSC group and 0.775 (0.398–1.441) in the non-rPSC group, with no significant difference between the two groups (Mann–Whitney *U* test, *P* = 0.847). ROC analysis demonstrated that the maximum log10-transformed antibody titer had poor ability to discriminate rPSC from non-rPSC, with an AUC of 0.531 (95% CI, 0.215–0.847) (data not shown).

Because pre-transplant antibody titers varied substantially among patients, we then evaluated fold changes relative to each patient’s pre-transplant baseline. FCearly, FC90, and FC180 did not differ significantly between the rPSC and non-rPSC groups (Mann–Whitney *U* test, *P* = 0.729, 0.526, and 0.452, respectively) (Supplementary Table S2). Because the timing of antibody re-elevation varied among patients, we next evaluated the maximum fold change observed during follow-up relative to the pre-transplant baseline (FCmax). The median (IQR) FCmax was 1.381 (0.898–3.863) in the rPSC group and 0.514 (0.320–0.748) in the non-rPSC group, representing a significant difference between the groups (Mann–Whitney *U* test, *P* = 0.041) (Fig. 3c). ROC analysis demonstrated that FCmax yielded an AUC of 0.781 (95% CI, 0.542–1.000) for discriminating rPSC (Fig. 3d). The optimal cutoff value determined using the Youden index was 0.996. Sensitivity analysis including the three patients with post-transplant changes in UC activity yielded similar results, with an AUC of 0.731 (95% CI, 0.506–0.956) for FCmax (data not shown). To further investigate the relationship between antibody re-elevation and the development of rPSC, we performed an exploratory longitudinal analysis using a fold-change threshold of >1.0, because the optimal cutoff derived from the ROC analysis (0.996) was essentially equivalent to the pre-transplant baseline. During follow-up, a first antibody re-elevation above the pre-transplant level was observed in seven patients. Of these, six (85.7%) were subsequently diagnosed with rPSC, whereas one remained free of recurrence throughout follow-up. Among the six patients who developed rPSC after the first antibody re-elevation, the median lead time from the first antibody re-elevation to the diagnosis of rPSC was 237.5 days (range, 42–1089 days) (Supplementary Table S3). Thirteen patients did not experience a first antibody re-elevation, and only two of them subsequently developed rPSC. Together, these findings indicate that post-transplant antibody re-elevation above the pre-transplant baseline is associated with rPSC development and may precede its diagnosis.

### Association Between Antibody Titers and Histopathological Findings

A total of 82 matched post-transplant liver biopsy–serum pairs were available from the entire cohort of 23 patients. After excluding all matched pairs derived from the three patients who experienced changes in UC disease activity during follow-up, 71 matched pairs from 20 patients remained for the association analysis (Fig. 1). The median interval between serum collection and liver biopsy was 0 days (range, −69 to 90 days). Two pathologists independently scored periductal fibrosis, neutrophilic and/or lymphocytic cholangitis, and periportal CK7-positive metaplastic cells. Separately, each biopsy specimen was independently assessed for histopathological findings of rPSC according to the 2024 diagnostic criteria. Because CK7-stained slides were unavailable for two biopsy specimens obtained at outside institutions from one patient, periportal CK7-positive metaplastic cells were evaluated in 69 specimens. Rejection Activity Index (RAI) scores were also available for 69 specimens. The weighted κ values for interobserver agreement were 0.600 (*P* = 8.1 × 10⁻⁸) for periductal fibrosis, 0.884 (*P* = 5.84 × 10⁻¹⁴) for neutrophilic and/or lymphocytic cholangitis, and 1.000 (*P* < 0.001) for periportal CK7-positive metaplastic cells (data not shown). Representative histopathological findings are shown in Fig. 4a–c.

**Fig. 4.**
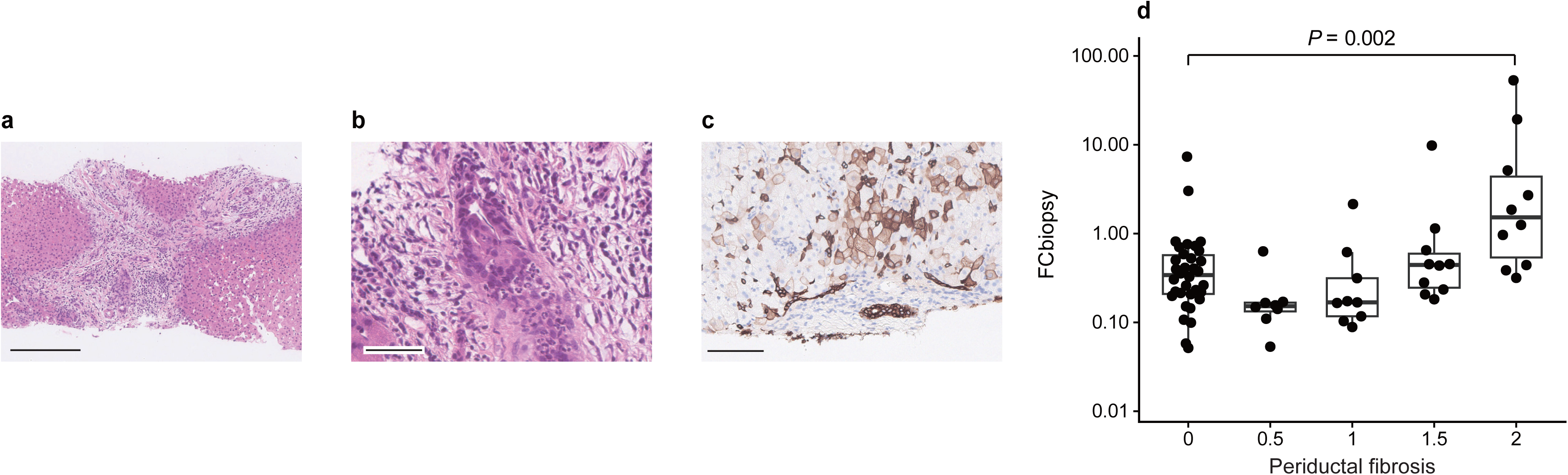
Representative histological findings and association of FCbiopsy with periductal fibrosis after liver transplantation. a–c Representative histopathological findings from biopsy specimens with a score of 2 (involvement of two or more portal tracts). a Periductal fibrosis (scale bar, 250 µm). b Neutrophilic and/or lymphocytic cholangitis (scale bar, 50 µm). c Periportal CK7-positive metaplastic cells (scale bar, 100 µm). d Distribution of FCbiopsy according to the periductal fibrosis score. Boxes represent the median and interquartile range, and dots represent individual biopsy specimens. Scores of 0.5 and 1.5 represent averaged discordant scores assigned by the two pathologists. The bracket and *P* value indicate the statistically significant comparison between scores 0 and 2; nonsignificant comparisons are not shown. FCbiopsy, fold change in antibody titer at the time of liver biopsy relative to the pre-transplant baseline

The relationships between histopathological scores and antibody titers were analyzed using linear mixed-effects models. Using log10-transformed antibody titers at the time of liver biopsy as the outcome variable, antibody titers differed significantly according to the periductal fibrosis score and the periportal CK7-positive metaplastic cell score, whereas no significant differences were observed according to neutrophilic and/or lymphocytic cholangitis or RAI score (Fig. 5a). The distributions of log10-transformed antibody titers across all histological severity scores are presented in Supplementary Fig. S2. The strongest association was observed for specimens assigned a periductal fibrosis score of 2, which exhibited significantly higher antibody titers than those assigned a score of 0 (β = 0.690, 95% CI 0.321–1.058, *P* < 0.001).

**Fig. 5.**
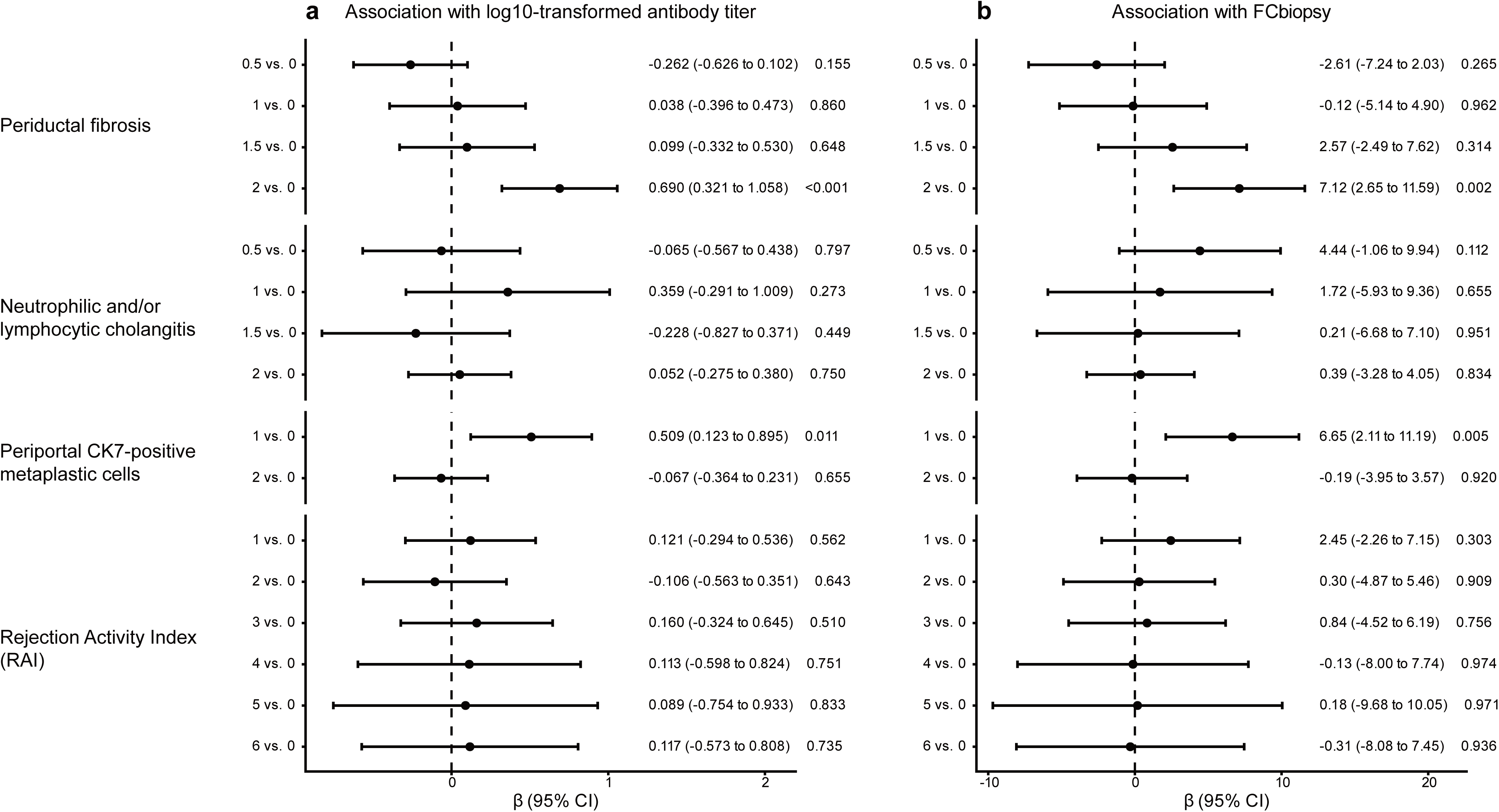
Forest plot of associations between anti-integrin αvβ6 autoantibody levels and histopathological findings. Forest plots showing the associations between histopathological scores and antibody measures. a Associations with log10-transformed anti-integrin αvβ6 autoantibody titers. b Associations with FCbiopsy. Estimates were obtained using linear mixed-effects models with patient ID as a random intercept. β coefficients, 95% confidence intervals, and P values are shown for each histopathological score compared with the reference category (score 0). Histopathological scores included periductal fibrosis, neutrophilic and/or lymphocytic cholangitis, periportal CK7-positive metaplastic cells, and Rejection Activity Index (RAI). FCbiopsy, fold change in antibody titer at the time of liver biopsy relative to the pre-transplant baseline

When FCbiopsy was used as the outcome variable, similar associations were observed (Fig. 5b). The distributions of FCbiopsy according to the scores for neutrophilic and/or lymphocytic cholangitis, periportal CK7-positive metaplastic cells, and RAI are presented in Supplementary Fig. S2, whereas the distribution according to the periductal fibrosis score is shown in Fig. 4d. Specimens assigned a periductal fibrosis score of 2 exhibited significantly higher FCbiopsy values than those assigned a score of 0 (β = 7.12, 95% CI 2.65–11.59, *P* = 0.002). FCbiopsy also differed according to the periportal CK7-positive metaplastic cell score, whereas no significant differences were observed according to neutrophilic and/or lymphocytic cholangitis or RAI score.

## Discussion

To our knowledge, this is the first study to comprehensively characterize longitudinal changes in anti-integrin αvβ6 autoantibody titers in adult patients after LT for PSC. Antibody titers decreased significantly at the first postoperative measurement, and subsequent re-elevation was observed in a considerable number of patients; notably, titers exceeded the individual pre-transplant baseline in most patients who developed rPSC. The initial postoperative decrease is consistent with previous observations in pediatric PSC [19] and likely reflects reduced antigenic stimulation after removal of the diseased liver. Antibody re-elevation above the individual baseline may reflect recurrence-associated immune reactivation rather than simple inter-individual differences in baseline antibody levels.

However, the relationship between antibody kinetics and histopathological findings was heterogeneous. In most patients, antibody re-elevation coincided with the development of characteristic histopathological changes. However, in Patient PSC11 (Supplementary Fig. S1), characteristic histopathological findings of rPSC became evident only after the antibody titer had peaked and subsequently declined. In addition, Patients PSC06 and PSC16 (Supplementary Fig. S1) developed rPSC despite antibody titers never exceeding their individual pre-transplant baseline, indicating that antibody kinetics do not consistently mirror histopathological changes.

After excluding patients with post-transplant changes in UC activity, we evaluated FCmax in addition to the maximum log10-transformed antibody titer to account for substantial inter-individual differences in baseline antibody titers; a previous post-transplant study had evaluated only absolute antibody titers [19]. FCmax showed better discrimination of rPSC (AUC 0.781 vs. 0.531) from non-rPSC, whereas analyses at the first postoperative measurement and postoperative days 90 and 180 were not discriminatory, indicating that antibody reactivation occurs at variable time points during follow-up. Accordingly, FCmax may be more suitable for longitudinal monitoring than for fixed-time-point assessment.

The optimal cutoff was close to 1.0, indicating that antibody titers exceeding the individual pre-transplant baseline may reflect recurrence-associated antibody reactivation. Consistent with this interpretation, our exploratory longitudinal analysis showed that 85.7% of patients who exceeded their individual pre-transplant antibody level were subsequently diagnosed with rPSC, with the initial antibody rise preceding clinical diagnosis of rPSC by a median of approximately 8 months. These findings suggest that postoperative antibody re-elevation above the individual baseline may serve as a predictive marker for rPSC.

Our histopathological analyses demonstrated that both log10-transformed antibody titers and FCbiopsy were higher in specimens assigned a periductal fibrosis score of 2 than in those assigned a score of 0. A similar, although non-monotonic, association was observed according to the periportal CK7-positive metaplastic cell score. Importantly, expressing antibody titers as fold changes relative to the pre-transplant antibody titer preserved these pathological associations, indicating that FCbiopsy retains pathological information while accounting for inter-individual differences in pre-transplant antibody titers. Together with its superior performance for identifying rPSC, these findings support the potential utility of FC-based antibody measurements as biomarkers for longitudinal monitoring after LT.

Integrin αvβ6 promotes TGF-β-mediated biliary fibrosis, and inhibition of integrin αvβ6 suppresses biliary fibrosis in experimental models [17]. In the present study, anti-integrin αvβ6 autoantibody levels were more strongly associated with periductal fibrosis than with cholangitis or allograft rejection. Although the biological significance of integrin αvβ6 and anti-integrin αvβ6 autoantibodies in PSC-associated fibrosis remains unclear, these findings suggest that anti-integrin αvβ6 autoantibodies may reflect fibrosis-associated biological processes. Periportal CK7-positive hepatocytes have been used as a histological indicator of chronic cholestasis in PSC [20]. The association with the periportal CK7-positive metaplastic cell score may reflect biliary epithelial remodeling. However, this association was non-monotonic, with significant differences observed only between scores 0 and 1, and its biological significance remains uncertain, warranting validation in larger independent cohorts.

All patients with increased UC activity after LT showed concomitant increases in anti-integrin αvβ6 autoantibody titers that decreased after UC treatment, including one patient who did not develop rPSC. Consistent with previous reports linking antibody titers to UC disease activity [12], these findings indicate that UC activity should be considered when interpreting anti-integrin αvβ6 autoantibodies as biomarkers of rPSC and suggest that intestinal immune activation may contribute to antibody production, consistent with the proposed gut–liver immune axis between UC and PSC.

This study has several limitations. First, this was a single-center retrospective study including only 23 patients, which may limit the generalizability of our findings. In addition, post-transplant UC activity was assessed retrospectively based on clinical records rather than by protocolized endoscopic evaluation, and subclinical changes in UC activity may therefore have been missed. Second, serum samples and liver biopsy specimens were collected as part of routine clinical practice, resulting in variable sampling intervals that precluded precise assessment of the relationship between antibody kinetics and rPSC or histopathological changes. Moreover, baseline serum samples were not uniformly collected immediately before transplantation, which may have introduced variability into fold-change measurements relative to the pre-transplant antibody titer. Finally, FCmax was defined as the maximum fold change observed during follow-up and should therefore be regarded as an exploratory longitudinal marker rather than a fixed-time-point biomarker. The optimal cutoff identified in this study should also be considered exploratory until validated in an independent cohort.

In conclusion, anti-integrin αvβ6 autoantibody titers decreased after liver transplantation. Subsequent re-elevation above the pre-transplant antibody titer was associated with the development of rPSC. These findings suggest that fold-change measurements relative to the pre-transplant antibody titer may serve as biomarkers for longitudinal monitoring of rPSC after LT. Future prospective validation in larger independent cohorts is warranted.

## Acknowledgements

We are deeply grateful to all the patients who provided serum samples for this study. We thank Shino Yamaguchi and Taichi Ito for their valuable technical assistance. Some of the anti-integrin αvβ6 ELISA kits used in this study were provided free of charge by Medical and Biological Laboratories Co., Ltd.; the remaining kits were purchased. Finally, we thank Editage for editing the English text.

## Author Contributions

Koki Chikugo contributed to the study design, data collection, data analysis and interpretation, and drafting of the manuscript. Takeshi Kuwada and Masahiro Shiokawa contributed to the conception and design of the study, interpretation of the data, supervision of the study, and critical revision of the manuscript. Yoshihiro Nishikawa, Hiroyuki Yoshida, Tatsuki Hirai, Nagomi Mankawa, Ryo Ito, Kota Hashimoto, Yuki Mori, Fumioki Toyoda, Ayako Hirata, Kenji Sawada, Takafumi Yanaidani, Masataka Yokode, Yuya Muramoto, Sakiko Ota, Tomonori Hirano, Yuko Sogabe, Nobuyuki Kakiuchi, and Tomoaki Matsumori contributed to data acquisition and interpretation and critically reviewed the manuscript. Yasuhide Takeuchi and Hironori Haga contributed to the histopathological assessment and interpretation of the pathological findings. Hajime Yamazaki contributed to the statistical analysis and interpretation of the data. Takashi Ito and Etsuro Hatano contributed to the acquisition and interpretation of clinical and surgical data related to liver transplantation. Tsutomu Chiba and Hiroshi Seno contributed to the interpretation of the data, supervision of the study, and critical revision of the manuscript. All authors reviewed and approved the final version of the manuscript.

## Data Availability Statement

The data that support the findings of this study are available from the corresponding author upon reasonable request.

---

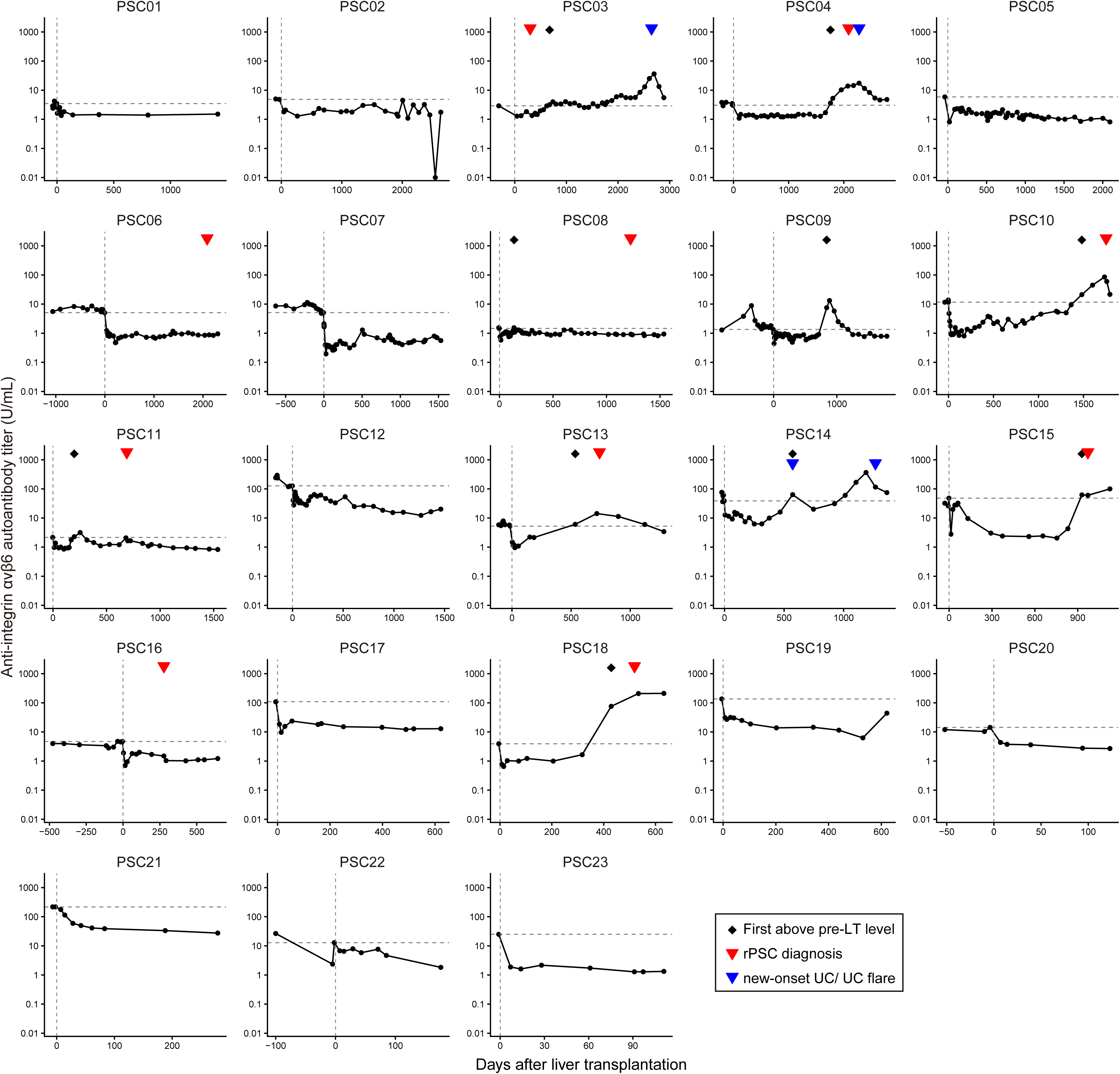

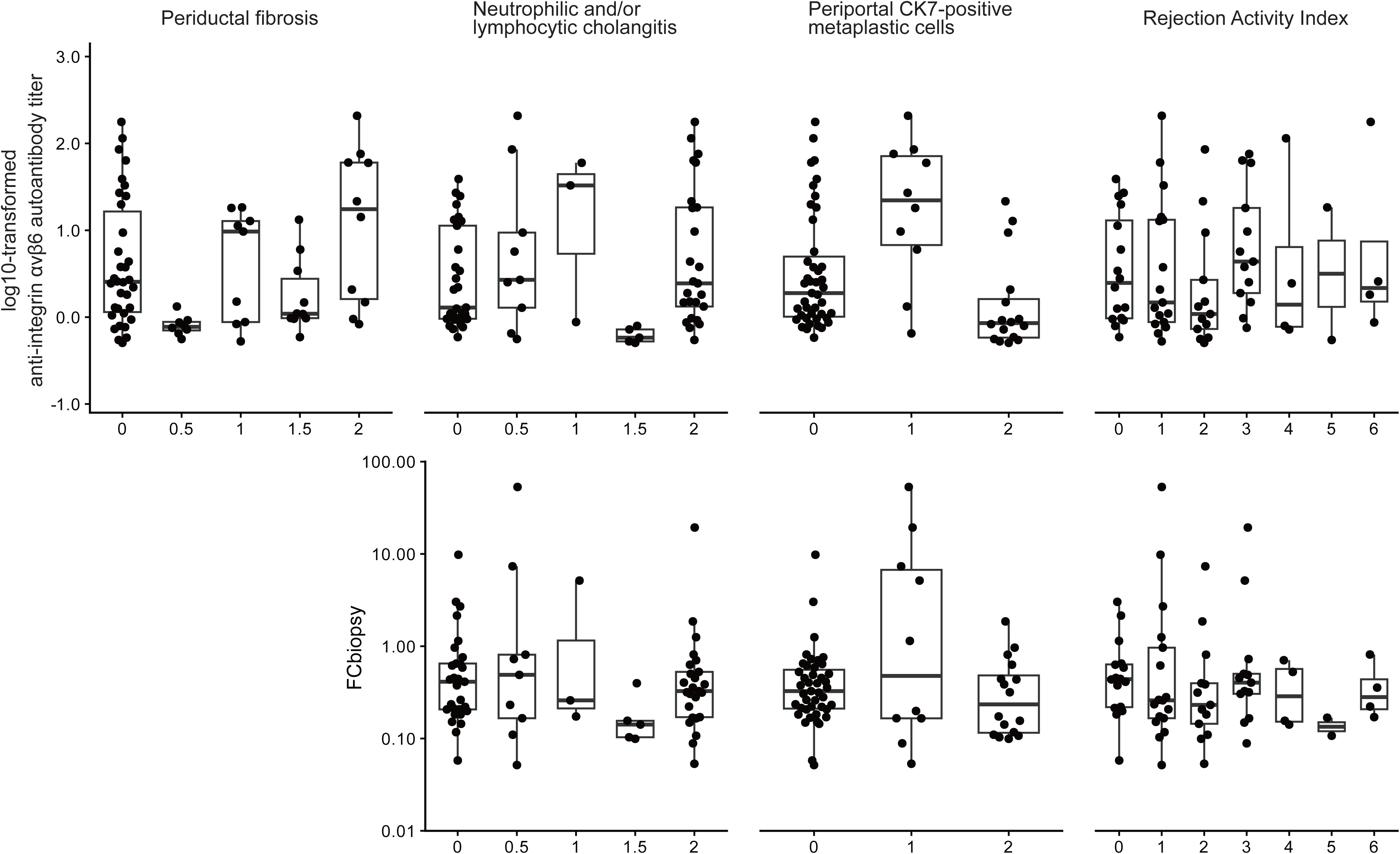

## References

1. Trivedi PJ, Bowlus CL, Yimam KK, et al. Epidemiology, Natural History, and Outcomes of Primary Sclerosing Cholangitis: A Systematic Review of Population-based Studies. Clin Gastroenterol Hepatol. 2022;20(8):1687–700 e4.

2. Tanaka A, Mori M, Matsumoto K, et al. Increase trend in the prevalence and male-to-female ratio of primary biliary cholangitis, autoimmune hepatitis, and primary sclerosing cholangitis in Japan. Hepatol Res. 2019;49(8):881–9.

3. Bowlus CL, Arrive L, Bergquist A, et al. AASLD practice guidance on primary sclerosing cholangitis and cholangiocarcinoma. Hepatology. 2023;77(2):659–702.

4. Vitor IC, Tome MR, Amador WFO, et al. Primary Sclerosing Cholangitis Recurrence After Liver Transplantation: A Systematic Review and Updated Meta-Analysis. Liver Int. 2026;46(6):e70673.

5. Ravikumar R, Tsochatzis E, Jose S, et al. Risk factors for recurrent primary sclerosing cholangitis after liver transplantation. J Hepatol. 2015;63(5):1139–46.

6. Akamatsu N, Hasegawa K, Egawa H, et al. Donor age (≥45 years) and reduced immunosuppression are associated with the recurrent primary sclerosing cholangitis after liver transplantation – a multicenter retrospective study. Transpl Int. 2021;34(5):916–29.

7. Naitoh I, Isayama H, Akamatsu N, et al. The 2024 diagnostic criteria for primary sclerosing cholangitis. J Gastroenterol. 2025;60(10):1221–31.

8. Graziadei IW, Wiesner RH, Batts KP, et al. Recurrence of primary sclerosing cholangitis following liver transplantation. Hepatology. 1999;29(4):1050–6.

9. Banff Working Group, Demetris AJ, Adeyi O, et al. Liver biopsy interpretation for causes of late liver allograft dysfunction. Hepatology. 2006;44(2):489–501.

10. Lazaridis KN, LaRusso NF. Primary Sclerosing Cholangitis. N Engl J Med. 2016;375(12):1161–70.

11. Dienes HP, Lohse AW, Gerken G, et al. Bile duct epithelia as target cells in primary biliary cirrhosis and primary sclerosing cholangitis. Virchows Arch. 1997;431(2):119–24.

12. Kuwada T, Shiokawa M, Kodama Y, et al. Identification of an Anti-Integrin alphavbeta6 Autoantibody in Patients With Ulcerative Colitis. Gastroenterology. 2021;160(7):2383–94 e21.

13. Muramoto Y, Nihira H, Shiokawa M, et al. Anti-Integrin alphavbeta6 Antibody as a Diagnostic Marker for Pediatric Patients With Ulcerative Colitis. Gastroenterology. 2022;163(4):1094–7 e14.

14. Yoshida H, Shiokawa M, Kuwada T, et al. Anti-integrin alphavbeta6 autoantibodies in patients with primary sclerosing cholangitis. J Gastroenterol. 2023;58(8):778–89.

15. Yasuda M, Shiokawa M, Kuwada T, et al. Anti-integrin alphavbeta6 autoantibody in primary sclerosing cholangitis: a Japanese nationwide study. J Gastroenterol. 2025;60(1):118–26.

16. Larjava H, Koivisto L, Hakkinen L, et al. Epithelial integrins with special reference to oral epithelia. J Dent Res. 2011;90(12):1367–76.

17. Koivisto L, Bi J, Hakkinen L, et al. Integrin alphavbeta6: Structure, function and role in health and disease. Int J Biochem Cell Biol. 2018;99:186–96.

18. Weil P, van den Bruck R, Ziegenhals T, et al. β6 integrinosis: a new lethal autosomal recessive ITGB6 disorder leading to impaired conformational transitions of the α(V)β6 integrin receptor. Gut. 2020;69(7):1359–61.

19. Maeda Y, Umetsu S, Hiejima E, et al. Anti-Integrin αvβ6 Autoantibodies as Diagnostic and Monitoring Biomarkers for Pediatric-Onset Primary Sclerosing Cholangitis. Hepatol Res. 2026; doi:10.1111/hepr.70211

20. Sjoblom N, Boyd S, Kautiainen H, et al. Novel histological scoring for predicting disease outcome in primary sclerosing cholangitis. Histopathology. 2022;81(2):192–204.

21. Yamashiki N, Haga H, Ueda Y, et al. Use of Nakanuma staging and cytokeratin 7 staining for diagnosing recurrent primary biliary cholangitis after living-donor liver transplantation. Hepatol Res. 2020;50(4):478–87.

22. Demetris AJ, Bellamy C, Hubscher SG, et al. 2016 Comprehensive Update of the Banff Working Group on Liver Allograft Pathology: Introduction of Antibody-Mediated Rejection. Am J Transplant. 2016;16(10):2816–35.

