## Supplementary Information for "Anti-integrin αvβ6 Autoantibodies in the Recurrence of Primary Sclerosing Cholangitis Following Liver Transplantation"

**Journal of Gastroenterology**

Koki Chikugo, Takeshi Kuwada*, Masahiro Shiokawa, Yoshihiro Nishikawa, Hiroyuki Yoshida, Tatsuki Hirai, Nagomi Mankawa, Ryo Ito, Kota Hashimoto, Yuki Mori, Fumioki Toyoda, Ayako Hirata, Kenji Sawada, Takafumi Yanaidani, Masataka Yokode, Yuya Muramoto, Sakiko Ota, Tomonori Hirano, Yuko Sogabe, Nobuyuki Kakiuchi, Tomoaki Matsumori, Yasuhide Takeuchi, Hajime Yamazaki, Takashi Ito, Tsutomu Chiba, Hironori Haga, Etsuro Hatano, and Hiroshi Seno

**Corresponding Author:** Takeshi Kuwada

Department of Gastroenterology and Hepatology, Kyoto University Graduate School of Medicine, Kyoto, Japan

54 Shogoin-kawahara-cho, Sakyo-ku, Kyoto 606-8507, Japan

**Supplementary Fig. S1 Longitudinal anti-integrin αvβ6 autoantibody titers in individual patients after liver transplantation**

Each panel represents one patient. The dashed vertical line indicates the time of liver transplantation (day 0). The dashed horizontal line indicates the individual pre-transplant antibody titer used as the baseline for fold-change calculations. Black diamonds indicate the time of the first postoperative antibody titer exceeding the pre-transplant level (first above pre-LT level). Red inverted triangles indicate the time of recurrent primary sclerosing cholangitis (rPSC) diagnosis. Blue inverted triangles indicate the onset of post-transplant ulcerative colitis (UC) or UC flare. Antibody titers are displayed on a logarithmic scale

**Supplementary Fig. S2 Distribution of anti-integrin αvβ6 autoantibody indices according to histological severity scores**

Distribution of log10-transformed anti-integrin αvβ6 autoantibody titers (upper panels) according to histological severity scores for periductal fibrosis, neutrophilic and/or lymphocytic cholangitis, periportal CK7-positive metaplastic cells, and Rejection Activity Index (RAI). FCbiopsy values (lower panels) are shown for neutrophilic and/or lymphocytic cholangitis, periportal CK7-positive metaplastic cells, and RAI; the corresponding analysis for periductal fibrosis is presented in Fig. 4d. Boxes represent the median and interquartile range, and dots represent individual biopsy specimens. Scores of 0.5 and 1.5 represent the average of discordant scores assigned independently by the two pathologists. FCbiopsy, fold change in antibody titer at the time of liver biopsy relative to the pre-transplant baseline

**Supplementary Table S1 Timing of serum sample collection and inclusion in study analyses**

Values indicate the timing of serum sample collection relative to LT. Pre-LT values represent days before LT, whereas first postoperative (Early post-LT) and postoperative day 90 (Post-LT day 90) values represent days after LT. The three-time-point antibody analysis included patients with serum samples available at all three time points. The rPSC/histopathology association analyses included 20 patients after excluding three patients with post-transplant changes in UC activity. “Yes” and “–” indicate inclusion and non-inclusion in the respective analysis. NA, not available; LT, liver transplantation; UC, ulcerative colitis.

| ID | Pre-LT (days before LT) | Early post-LT (days after LT) | Post-LT day 90 (days after LT) | Three-time-point antibody analysis | rPSC/histopathology association analyses |
|---|---|---|---|---|---|
| PSC01 | 2 | 3 | 62 | Yes | Yes |
| PSC02 | 31 | 41 | 69 | Yes | Yes |
| PSC03 | 310 | 42 | NA | - | - |
| PSC04 | 15 | 103 | 103 | Yes | - |
| PSC05 | 39 | 21 | 88 | Yes | Yes |
| PSC06 | 5 | 35 | 90 | Yes | Yes |
| PSC07 | 5 | 1 | 100 | Yes | Yes |
| PSC08 | 2 | 5 | 87 | Yes | Yes |

| ID | Pre-LT (days before LT) | Early post-LT (days after LT) | Post-LT day 90 (days after LT) | Three-time-point antibody analysis | rPSC/histopathology association analyses |
|---|---|---|---|---|---|
| PSC09 | 6 | 1 | 92 | Yes | Yes |
| PSC10 | 1 | 6 | 97 | Yes | Yes |
| PSC11 | 3 | 11 | 101 | Yes | Yes |
| PSC12 | 2 | 6 | 89 | Yes | Yes |
| PSC13 | 16 | 3 | NA | - | Yes |
| PSC14 | 2 | 5 | 85 | Yes | - |
| PSC15 | 1 | 13 | 62 | Yes | Yes |
| PSC16 | 4 | 3 | 90 | Yes | Yes |
| PSC17 | 6 | 7 | NA | - | Yes |
| PSC18 | 5 | 7 | 104 | Yes | Yes |
| PSC19 | 5 | 7 | 104 | Yes | Yes |
| PSC20 | 4 | 7 | 94 | Yes | Yes |
| PSC21 | 2 | 7 | 83 | Yes | Yes |
| PSC22 | 2 | 7 | 85 | Yes | Yes |
| PSC23 | 1 | 7 | 91 | Yes | Yes |

**Supplementary Table S2 Comparison of fold changes in anti-integrin αvβ6 autoantibody titers at predefined postoperative time points between patients with and without rPSC**

Fold changes relative to the pre-transplant baseline antibody titer (FCearly, FC90, and FC180) were compared between patients with and without rPSC. FCearly was calculated using the first postoperative serum sample. FC90 and FC180 were calculated using the serum samples obtained closest to postoperative days 90 and 180, respectively, among those collected within ±30 days of each time point. Data are presented as median (interquartile range). *P* values were calculated using the Mann–Whitney *U* test.

| Variable | non-rPSC | rPSC | *P* value |
|---|---|---|---|
| FCearly | 0.348 (0.213–0.478) | 0.342 (0.230–0.422) | 0.729 |
| FC90 | 0.259 (0.159–0.403) | 0.368 (0.234–0.458) | 0.526 |
| FC180 | 0.166 (0.132–0.417) | 0.360 (0.207–0.616) | 0.452 |

**Supplementary Table S3 Exploratory longitudinal analysis of postoperative antibody re-elevation above the pre-transplant baseline**

Postoperative antibody re-elevation was defined as the first antibody re-elevation above the individual pre-transplant antibody level (fold change >1.0). The exploratory analysis was performed in the association analysis cohort. Lead time was defined as the interval from the first antibody re-elevation above the pre-transplant level to the diagnosis of rPSC. PSC, primary sclerosing cholangitis; rPSC, recurrent primary sclerosing cholangitis.

| Parameter | Value |
| --- | --- |
| Patients included in the association analysis | 20 |
| Patients experiencing a first antibody re-elevation above the pre-transplant level, n (%) | 7 (35%) |
| Outcome among patients experiencing a first antibody re-elevation above the pre-transplant level (n = 7) | |
| Subsequently diagnosed with rPSC, n (%) | 6 (85.7%) |
| Remained recurrence-free throughout follow-up, n (%) | 1 (14.3%) |
| Lead time from first antibody re-elevation above the pre-transplant level to rPSC diagnosis, median days (range) | 237.5 (42–1089) |
